# Disparities in Lung Cancer Screening Utilization at Two Health Systems in the Southeastern US

**DOI:** 10.1101/2024.05.12.24307248

**Authors:** Soumya J Niranjan, Desiree Rivers, Rekha Ramachandran, J Edward Murrell, Kayleigh C Curry, Mohammed Mubasher, Eric Flenaugh, Mark T Dransfield, Sejong Bae, Isabel C Scarinci

**Author notes:** **Corresponding author: Soumya J Niranjan**, University of Alabama at Birmingham, **United States.**.

## Abstract

**Purpose:** Low-dose computed tomography lung cancer screening is effective for reducing lung cancer mortality. It is critical to understand the lung cancer screening practices for screen-eligible individuals living in Alabama and Georgia where lung cancer is the leading cause of cancer death. High lung cancer incidence and mortality rates are attributed to high smoking rates among underserved, low income, and rural populations. Therefore, the purpose of this study: (1) to define sociodemographic and clinical characteristics of patients who were screened for lung cancer at an Academic Medical Center (AMC) in Alabama and a Safety Net Hospital (SNH) in Georgia.

**Methods:** A retrospective cohort study of patient electronic health records who received lung cancer screening between 2015 to 2020 was performed to identify the study population and outcome variable measures. Chi-square tests and Student t-tests were used to compare screening uptake across patient demographic and clinical variables. Bivariate and multivariate logistic regressions determined significant predictors of lung cancer screening uptake.

**Results:** At the AMC, 67,355 were identified as eligible for LCS and 1,129 were screened. In bivariate analyses, there were several differences between those who were screened and those who were not screened. Screening status in the site at Alabama varied significantly by age (P<0.01), race (P<0.001), marital status (P<0.01), smoking status (P<0.01) health insurance (P<0.01), median income (P<0.01), urban status (P<0.01) and distance from UAB (P<0.01). Those who were screened were more likely to have lesser comorbidities (2.31 vs. 2.53; P<0.001). At the SNH, 11,011 individuals were identified as screen-eligible and 500 were screened. In the site at Georgia, screening status varied significantly by race (P<0.01), health insurance (P<0.01), and distance from site (P<0.01). At the AMC, the odds of being screened increased significantly if the individual was a current smoker compared to former smoker (OR=3.21; P<0.01). At the SNH, the odds of being screened for lung cancer increased significantly with every unit increase in co-morbidity count (OR = 1.12; P=0.01)

**Conclusion:** The study provides evidence that LCS has not reached all subgroups and that additional targeted efforts are needed to increase lung cancer screening uptake. Furthermore disparity was noticed between adults living closer to screening institutions and those who lived farther.

## INTRODUCTION

Lung cancer is the leading cause of cancer-related mortality in the United States.^1^ After tobacco cessation, one of the most effective mechanisms for reducing lung cancer mortality is through annual lung cancer screening (LCS) with low-dose computed tomography (LDCT). ^2–4^ The National Lung Screening Trial (NLST) showed that annual screening of high-risk individuals with LDCT offered a 20% relative decrease in lung cancer mortality and a 6.7% reduction in all-cause mortality when compared with chest radiography. ^4^ Based on these results, since December 2013, the US Preventive Services Task Force (USPSTF) has recommended LCS for high-risk individuals (those 55-80 years of age who currently smoke cigarettes or quit within the past 15 years, with at least 30 pack-years total smoking history).^5^ It is to be noted that eligibility has since been revised to include those above 50 yrs. and smoking history of at least 20 pack-years. ^6^ Unfortunately, nearly 10 years after the guidelines released, LCS utilization is woefully low, at <6%.^7^ Screening rates varied geographically, with higher rates in several Northeastern states(∼15.2%) that have lower lung cancer burden and lower rates in several Southern states (<5%) that have high lung cancer burden.^7^

Specifically, the percentage of eligible individuals screened for lung cancer in Alabama and Georgia is less than the national average at 5%^7^ . In both states, high lung cancer incidence and mortality rates are attributed to high smoking rates among underserved, low income, and rural populations. ^7^ The need to address these disparities is high since both these states have more than twice the Black population as the national average (26.8% vs 13.4%) and Black individuals are at higher risk of dying from lung cancer.^8^ Black individuals in these states are also more likely to be uninsured than Whites (12% vs 8.8%), and more likely to be covered by Medicaid (31.5% vs 15%). Alabama is currently the only state that that does not cover lung cancer screening among Medicaid recipients, which impacts access. Rurality is another likely contributor to Alabama’s and Georgia’s increased lung cancer mortality since 82% and 77% of its counties are classified as rural. ^9^ Compared to its urban counterparts, rural populations have increased rates of lung cancer^10,11^ and face unique barriers including limited access to healthcare, provider shortages, higher poverty, and lack of health insurance coverage.^12^ In addition to rurality, distance to travel for screening is a major factor in determining likelihood of screening.^13^ Previous studies have determined associations between common rural factors such as poverty / distance to travel for screening and lower screening participation, making this an important area for potential intervention.^14,15,16^

Therefore, it is critical to better understand the current screening landscape in these at-risk populations in the Southeastern US.^17^. It will facilitate targeted interventions that will increase uptake, potentially improve survival while paying particular attention to inequalities in screening since it ultimately has the potential to exacerbate inequalities in lung cancer survival. ^18,19,20^ Moreover, although individual institutions ^21,22^ have described LCS implementation strategies and outcomes in single center analyses, there are limited studies^23^ that integrate cohorts. Therefore, the purpose of this study: (1) to define sociodemographic and clinical characteristics of patients who were screened for lung cancer at an Academic Medical Center (AMC) in Alabama and a Safety Net Hospital (SNH) in Georgia. We hypothesize that screening has not permeated all patient subpopulations.

## METHODS & MATERIALS

### Data Source

In order to identify the study population and outcome variable measures, patient-level data was obtained from each of the institutions’ electronic health records. This retrospective cohort study was performed with approval from the Institutional Review Board at the University of Alabama at Birmingham and Grady Health Systems which is a major affiliate of Morehouse School of Medicine (MSM). Written informed consent was waived because of the retrospective nature of the study in accordance with institution policies.

### Study population

We identified a cohort of patients who received lung cancer screening between 2015 to 2020. For the same years, we identified [via i2b2 (Informatics for Integrating Biology and the Bedside) – used for cohort identification / denominator] screen-eligible patients based on the USPSTF guidelines at the time the study^5^: (i) between 55 and 80 years; and (ii) smoking history of at least 30 pack-years and currently smoke or have quit within the past 15 years and (iii) no previous diagnoses of lung cancer. It is to be noted that eligibility has since been revised to include those above 50 yrs. and smoking history of at least 20 pack-years.^6^

### Measures and variables of interest

The primary outcome, dependent variable, of interest was patient completion of LCS. A dichotomous variable was created for LCS status where patients were considered “screened” when LCS using LDCT was completed and “not screened” otherwise. Patient demographic variables included in the analysis: Age ; Marital Status(grouped as Married/Living together; Separated/divorced/widowed; Single, never married) Sex (female; Male); Race/ethnicity Health Insurance (grouped as Private; Public ; combination of Public and Private and No Insurance); Income; Rurality (using Rural–Urban Continuum Codes [RUCCs]). ^24^ We categorize RUCC 1-3 as urban and 4-9 as rural. Income was categorized as <25K, 25K - <50K, 50K - <75K, ≥75K. Sex was measured as a dichotomous variable. Race/ethnicity consisted of 3 categories: White (referent), Black or African American and Other – which included Asian, Native Hawaiian/Pacific Islanders, Hispanic American Indian or Alaska Native, Co-morbidities: a comorbidity domain count was used to account for other related pathologies—the measure estimates one or more clinical conditions the patient was diagnosed with between 2015 and 2020. The variable included over 15,000 International Classification of Diseases, Ninth Revision, Clinical Modification (ICD-9-CM) diagnosis codes that appeared in the electronic records, aggregated into 202 clinically meaningful “conditions,” and then grouped into 17 broad domains. ^25^ We used patient’s residential zip code to estimate distance to their respective medical facility (AMC and SNH).

### Statistical analysis

Chi-square tests and Student t-tests were used to compare screening uptake across patient demographic and clinical variables. We used bivariate and multivariate logistic regressions to determine significant predictors of lung cancer screening uptake at UAB and Grady Hospital. Multivariate logistic regression analyses were conducted to examine the relationship between various predictor variables, including Hospital, Race, Gender, Marital Status, Smoking Status, RUCC, Distance from hospital, ACS Median Income, age, and Total Comorbidities, and the status of lung cancer screening. The threshold for statistical significance was set at the 0.05 level. We tested for LCS differences interactions with current smoking, race, and other variables by hospital. Based on the results of the significant interaction (p < 0.1), we examined the LCS relationship stratified by hospital (data not shown) to grasp the nature and direction of the relationships between variables. Statistical analysis was conducted using SAS version 9.4.

## RESULTS

### Descriptive characteristics of the patient cohort at UAB

Of the 67,355 identified as eligible for LCS, 1,129 were screened (1.67%) (Table 1). Of those 1,129 individuals screened, the mean age was 67.02, predominantly male (54.92%) and Non-Hispanic White (57.77%). A majority (67.05%) of those screened held a combination of public and private health insurance; were current smokers (53.95%) and lived in urban areas (87.95%)

**Table 1.** Patient Characteristics and Lung Cancer Screening at AMC

|  | **Total**<br>**(N = 67355)** | **Unscreened**<br>**(N =66226 )** | **Screened**<br>**(N =1129 )** | **p value** |
| --- | --- | --- | --- | --- |
| **Age (Mean (SD))** | 69.30 (7.45) | 69.34 (7.47) | 67.02 (5.97) | <0.01 |
| **Race** |  |  |  |  |
| African American | 16228 (25.02) | 15795 (24.77) | 433 (38.9) | <0.01 |
| Other | 1914 (2.95) | 1877 (2.94) | 37 (3.32) |  |
| White | 46727 (72.03) | 46084 (72.28) | 643 (57.77) |  |
| **Sex** |  |  |  |  |
| Female | 29513 (43.82) | 29004 (43.80) | 509 (45.08) | 0.39 |
| Male | 37842 (56.18) | 37222 (56.20) | 620 (54.92) |  |
| **Marital status** |  |  |  |  |
| Married/Living together | 36139 (56.95) | 35635 (57.15) | 504 (45.69) | <0.01 |
| Separated/divorced/widowed | 15777 (24.86) | 15466 (24.81) | 311 (28.20) |  |
| Single, never married | 11537(18.18) | 11249 (18.04) | 288 (26.11) |  |
| **Insurance status** |  |  |  |  |
| No Insurance | 383 (0.57) | 381 (0.58) | 2 (0.18) | <0.01 |
| Public | 36375 (54) | 36238 (54.72) | 137 (12.13) |  |
| Private/Public | 12630 (18.75) | 11873 (17.93) | 757 (67.05) |  |
| Private | 17366 (25.78) | 17133 (25.87) | 233 (20.64) |  |
| **Smoking status** |  |  |  |  |
| Current | 18343 (27.24) | 17742 (26.79) | 601 (53.95) | <0.01 |
| Former | 46505 (69.06) | 46020 (69.49) | 485 (43.54) |  |
| Never | 2321 (3.45) | 2296 (3.47) | 25 (2.24) |  |
| **ACS Median Income** |  |  |  |  |
| 25K - <50K | 29278 (44.07) | 28715 (43.96) | 563 (50.44) | <0.01 |
| 50K - <75K | 27074 (40.76) | 26700 (40.88) | 374 (33.51) |  |
| ≥75K | 10073 (15.16) | 9894 (15.15) | 179 (16.04) |  |
| **Urban status (based on RUCC 2013 )** |  |  |  |  |
| Rural | 18837 (28) | 18701 (28.27) | 136 (12.05) | <0.01 |
| Urban | 48435 (72) | 47442 (71.73) | 993 (87.95) |  |
| **Comorbidities** |  |  |  |  |
| AMI and MI | 9500 (14.1) | 9291 (14.03) | 209 (18.51) | <0.01 |
| CVD | 13289 (19.73) | 12994 (19.62) | 295 (26.13) | <0.01 |
| CHF | 12409 (18.42) | 12138 (18.33) | 271 (24) | <0.01 |
| CKD | 11167 (16.58) | 10931 (16.51) | 236 (20.9) | <0.01 |
| CVA/Stroke | 6461 (9.59) | 6313 (9.53) | 148 (13.11) | <0.01 |
| Dementia | 3079 (4.57) | 3041 (4.59) | 38 (3.37) | 0.05 |
| Diabetes | 19829 (29.44) | 19409 (29.31) | 420 (37.2) | <0.01 |
| ESRD | 2796 (4.15) | 2765 (4.18) | 31 (2.75) | 0.02 |
| Hemi Paraplegia | 2563 (3.81) | 2513 (3.79) | 50 (4.43) | 0.27 |
| HIV/AIDS | 858 (1.27) | 799 (1.21) | 59 (5.23) | <0.01 |
| Liver Disease | 13365 (19.84) | 13058 (19.72) | 307 (27.19) | <0.01 |
| Malignancy | 18110 (26.89) | 17839 (26.94) | 271 (24) | 0.03 |
| PUD | 2556 (3.79) | 2487 (3.76) | 69 (6.11) | <0.01 |
| PVD | 16627 (24.69) | 16119 (24.34) | 508 (45) | <0.01 |
| **Total Comorbidities (Mean (SD)))** | 1.97 (1.96) | 1.96 (1.96) | 2.58 (2.15) | <0.01 |
| **Charlston Commorbidity index (Mean (SD))** | 2.53 (0.80) | 2.53 (0.80) | 2.31 (0.65) | <0.01 |
| **Distance from hospital** | 50.03 (47.93) | 50.51 (48.05) | 22.06 (28.67) | <0.01 |

*Urban: contains dual zip codes*

### Descriptive characteristics of the patient cohort at Grady Hospital

Of the 11,011 screen-eligible individuals, 500 were screened (4.5%). Of those individuals screened, mean age was 67.92 years, were predominantly Black (83.77%) and male (55.4%). Screened individuals were predominantly current smokers (98%); majority held public health insurance (78%) and lived in urban areas (99.8%) (Table 2).

**Table 2.** Patient Characteristics and Lung Cancer Screening at SNH

|  | **Total (N = 11011)** | **Unscreened**<br>**(N = 10511)** | **Screened**<br>**(N = 500)** | **p value** |
| --- | --- | --- | --- | --- |
| **Age (Mean (SD))** | 66.45 (7.50) | 66.41 (7.58) | 67.29 (5.52 ) | <0.01 |
| **Race** |  |  |  |  |
| African American | 8147 (75.71) | 7729 (75.32) | 418 (83.77) | <0.01 |
| Other | 513 (4.77) | 494 (4.81) | 19 (3.81) |  |
| White | 2101 (19.52) | 2039 (19.87) | 62 (12.42) |  |
| **Sex** |  |  |  |  |
| Female | 4354 (39.64) | 4131 (39.4) | 223 (44.6) | 0.02 |
| Male | 6630 (60.36) | 6353 (60.6) | 277 (55.4) |  |
| **Marital status** |  |  |  |  |
| Married/Living together | 1813 (17.25) | 1735 (17.32) | 78 (15.73) | <0.01 |
| Separated/divorced/widowed | 3318 (31.57) | 3130 (31.25) | 188 (37.9) |  |
| Single, never married | 5380 (51.18) | 5150 (51.42) | 230 (46.37) |  |
| **Insurance status** |  |  |  |  |
| Public | 6465 (58.71) | 6075 (57.8) | 390 (78) | <0.01 |
| Private/Public | - | - | - |  |
| Private | 1384 (12.57) | 1358 (12.92) | 26 (5.2) |  |
| Other | 3162 (28.72) | 3078 (29.28) | 84 (16.8) |  |
| **Smoking status** |  |  |  |  |
| Current | 10245 (93.04) | 9781 (93.05) | 464 (92.8) | 0.83 |
| Former | 766 (6.96) | 730 (6.95) | 36 (7.2) |  |
| **ACS Median Income** |  |  |  |  |
| <50K | 4356 (39.87) | 4158 (39.86) | 198 (40.08) | 0.77 |
| 50K - <75K | 4696 (42.98) | 4479 (42.94) | 217 (43.93) |  |
| ≥75K | 1874 (17.15) | 1795 (17.21) | 79 (15.99) |  |
| **Urban status (based on RUCC 2013 )** |  |  |  |  |
| Rural /Rural and Urban | 333 (3.03) | 332 (3.16) | 1 (0.2) | <0.01 |
| Urban | 10674 (96.97) | 10175 (96.84) | 499 (99.8) |  |
| **Comorbidities** |  |  |  |  |
| **AMI and MI** | 140 (1.27) | 128 (1.22) | 12 (2.4) | 0.02 |
| CeVD | 1069 (9.71) | 1022 (9.72) | 47 (9.4) | 0.81 |
| CHF | 635 (5.77) | 593 (5.64) | 42 (8.4) | <0.01 |
| CKD | 514 (4.67) | 472 (4.49) | 42 (8.4) | <0.01 |
| CVA/Stroke | 469 (4.26) | 440 (4.19) | 29 (5.8) | 0.08 |
| Dementia | 106 (0.96) | 98 (0.93) | 8 (1.6) | 0.14 |
| Diabetes | 1816 (16.49) | 1676 (15.95) | 140 (28) | <0.01 |
| ESRD | 93 (0.84) | 89 (0.85) | 4 (0.8) | 0.91 |
| Hemi Paraplegia | 2 (0.02) | 2 (0.02) | 0 (0) | >0.99 |
| HIV/AIDS | 543 (4.93) | 517 (4.92) | 26 (5.2) | 0.78 |
| Liver Disease | 113 (1.03) | 104 (0.99) | 9 (1.8) | 0.08 |
| Malignancy | 808 (7.34) | 768 (7.31) | 40 (8) | 0.56 |
| PUD | 133 (1.21) | 126 (1.2) | 7 (1.4) | 0.69 |
| PVD | 304 (2.76) | 273 (2.6) | 31 (6.2) | <0.01 |
| **Total Comorbidities (Mean (SD)))** | 0.61 (0.92) | 0.60 (0.91) | 0.87 (1.04) | <0.01 |
| **Distance from hospital** | 13.50 (19.96) | 13.72 (20.31 ) | 8.79 (9.04 ) | <0.01 |

*Urban: contains dual zip codes*

### Variables associated with Lung Cancer Screening at Academic Medical Center

In bivariate analyses, there were several differences between those who were screened and those who were not screened. Screened patients were significantly older than their unscreened counterparts (P<0.01), were more likely to be White (P<0.01), were more likely to be married (P<0.01), were more likely to be current smokers (P<0.01) were more likely to be insured (P<0.01), were more likely to have an income between 25K and 50K (P<0.01), more likely to live in urban areas (P<0.01) and more likely to reside closer to the AMC (P<0.01). Those who were screened were more likely to have lesser comorbidities (2.31 vs. 2.53; P<0.01). (Table 1).

When controlling for relevant variables, there were several significant variables associated with screening (Table 3). The odds of being screened increased significantly if the individual was a current smoker compared to former smoker (OR=3.21; P<0.01) and with increase in co-morbidity count (OR = 1.08; P<0.01). The odds of being screened decreased significantly with every additional 10-mile distance from UAB (OR= 0.85; P<0.01) and with every 10-year increase in patient age (OR=0.70; P<0.01). Individuals with a combination of public and private insurance were 5 times more likely to be screened for lung cancer (OR= 5.4; P<0.01). Compared to those with income more than $75K, individuals with an income between $50K and $75K were less likely to be screened for lung cancer (OR=0.80 ; P=0.03)

**Table 3.** Multivariable analysis of LCS at UAB

| Effect | Odds Ratio (95% CI) | P-value |
| --- | --- | --- |
| **RUCC 2013 Urban/rural** |  |  |
| Urban vs Rural/Urban and Rural | 1.028 (0.801, 1.318) | 0.83 |
| **Race** |  | 0.12 (type3) |
| African American vs White | 0.894 (0.754, 1.06) | 0.20 |
| Other vs White | 0.687 (0.466, 1.011) | 0.06 |
| **Sex** |  |  |
| Female vs Male | 0.95 (0.83, 1.088) | 0.46 |
| **Marital Status** |  | 0.49 (type3) |
| Married/Living together vs Single, never married | 0.945 (0.794, 1.125) | 0.53 |
| Separated/divorced/widowed vs Single, never married | 0.893 (0.742, 1.075) | 0.23 |
| **Smoking Status** |  |  |
| Current vs Former | 3.208 (2.795, 3.682) | <0.01 |
| **Distance from hospital (10 units increase)** | 0.848 (0.821, 0.876) | <0.01 |
| **Age (10 units increase)** | 0.704 (0.63, 0.786) | <0.01 |
| **Total Comorbidities (1 Unit increase)** | 1.075 (1.041, 1.109) | <0.01 |
| **Income based on ACS** |  | 0.15 (type3) |
| <25k vs ≥75k | 0.773 (0.436, 1.372) | 0.38 |
| 25k - <50K vs ≥75k | 0.893 (0.719, 1.11) | 0.31 |
| 50k - <75k vs ≥75k | 0.8 (0.653, 0.981) | 0.03 |
| **Insurance** |  | <0.01 (type3) |
| Private/Public vs Private | 5.403 (4.505, 6.48) | <0.01 |
| Public vs Private | 0.393 (0.307, 0.501) | <0.01 |

*Urban: contains dual zip codes*

### Variables associated with Lung Cancer Screening at Safety net hospital

Screened patients were more likely to be Black/African American by race (P<0.01), were more likely to hold public insurance (P<0.01), and were more likely to reside closer to the SNH (P<0.01). Controlling for relevant confounders, we found that that the odds of being screened for lung cancer increased significantly with every unit increase in co-morbidity count (OR = 1.12; P=0.01). In the model that included insurance and total co-morbidities (Table 4), patients residing in the urban areas were 4 times more likely to be screened -although not statistically significant (OR= 4.78; P = 0.14).

**Table 4.**
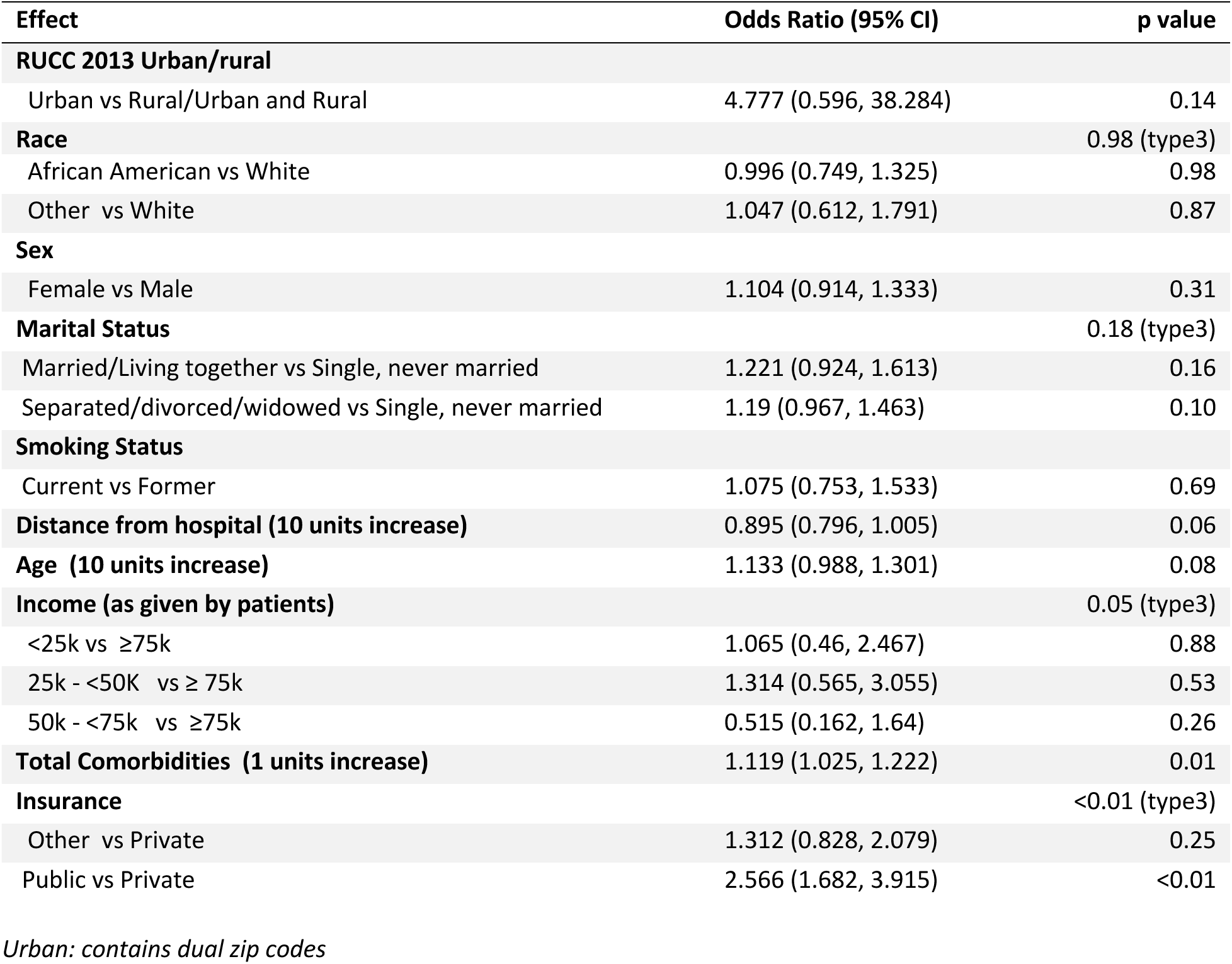
Multivariable analysis of LCS at Grady Hospital

## DISCUSSION

This study characterized individuals who have undergone LCS at two distinct health systems facilitated by a unique collaboration between institutions serving underserved health disparity populations.

At the AMC, we found 1,129 patients were screened for lung cancer between 2015 to 2020. These low screening numbers may in part be due to a decentralized program-which studies have shown to be inferior to centralized programs both in terms of screening rates and screening adherence after the first screening.^26,27^ This in turn leads to most eligible adults being reliant on opportunistic screening that present with multiple barriers.^28^ Lower screening numbers may also indicative of limited PCP knowledge of LCS guidelines as the point of first contact with the healthcare system for many patients, providers play a significant role in recommending lung cancer screening tests to and/or interpreting its results for patients.^29,30,31,32^ It was outside the scope of this study to explore physician knowledge and awareness and future studies should address these associations.

Equitable cancer screening is a critical part of reducing health disparities. Black individuals, while not statistically significant, were less likely to be screened for lung cancer at both sites. This study finding is corroborated by other studies conducted at single centers demonstrating racial disparities in LCS rates. ^17,22,33,34^ ^35^ This merits urgent attention since LCS may offer a greater mortality benefit for Black individuals than White individuals^36^, and non-Hispanic Black individuals are at higher risk of dying from lung cancer.^8^

At the AMC, patients who had a combination of public and private insurance were more likely to be screened when compared to those with private insurance only. Also, when compared to private insurance, those with public insurance were less likely to be screened. This is noteworthy since current tobacco use is highest among the uninsured and those with Medicaid compared to those with other public or private insurance^37^ and Alabama is 1 of 6 states to not cover lung cancer screening for Medicaid recipients both of which make access to screening difficult for those at greatest risk for lung cancer.^37^ This is in direct contrast to those screened at the SNH where a majority held public insurance (Medicare). Medicare provides a free Annual Wellness Visit that help ensure patients are up-to-date with recommended screenings, make a personalized screening schedule and patients may be more willing to be screened if they think subsequent procedures and treatment will be available and affordable.^38,39^ In this population with significantly more co-morbidities, healthcare disparities can be better addressed with coordinated services and continuity of care.

At both institutions, individuals reported as current smokers were more likely to be screened for lung cancer at both institutions. This has clinical implications since final coverage requires that screening-eligible beneficiaries who are current smokers receive smoking cessation counseling and information regarding tobacco cessation interventions.^40^ The results of the current study indicate that the screening-eligible population will have higher smoking rates, thus creating a substantial demand for cessation services and greater efforts are needed to integrate cessation services into clinical practice, particularly given that physician involvement can play a critical role in smoking cessation. ^41^ However, this finding should be interpreted with caution because medical records do not consistently capture tobacco use.

Distance to the institutions was a factor associated with lung cancer screening-odds of being screened reduced with increase in distance at both institutions. This is notable since we^42^ and others^43,44^ have demonstrated that geographical access to screening centers outside of urban medical centers are needed to improve uptake of lung cancer screening.

Our study has several limitations. As with all retrospective studies, our data reveal associations but does not provide evidence for causation. We report lung cancer screening rates of one academic medical center and a safety-net hospital which limits the generalizability of this study. Additionally, we recognize that the individuals who may be considered eligible is an overestimate, however we are unable to determine the number of screen-eligible individuals due to inadequate, inconsistent and unreliable documentation of tobacco use in medical records for calculating pack-years.^45,46^ Additionally, individuals with missing smoking history were not included in the analysis. Moreover, inaccuracies in reporting or incomplete documentation of smoking histories can prevent screen-eligible individuals who have access to care from being referred for LCS. ^47^ Despite these limitations, LCS patients described in this study are characterized based as a percentage of USPSTF-eligible individuals being seen at these institutions and has greater immediate impact in targeting sub-populations at risk for provision of LCS resources.

Specific targets for interventions at both sites can include engaging and empowering screen-eligible patients regarding lung cancer screening.^47–49^ in order to increase lung cancer screening rates. Furthermore, collection of smoking history in the EHR needs to be improved at both sites for increasing lung cancer screening since missing, outdated, and inaccurate smoking data in the EHR are mutable factors in identifying patients for lung cancer screening.^50^

## CONCLUSION

Lung cancer screening patients described in this study are characterized based as a percentage of USPSTF-eligible individuals being seen at these institutions and has greater immediate impact in targeting sub-populations at risk for provision of LCS resources. Findings from our study shows that there are overlapping factors between those seen at an AMC and a SNH that were associated with lung cancer screening. The study provides evidence that LCS has not reached all subgroups and that additional targeted efforts are needed to increase lung cancer screening uptake.

Furthermore, we saw a substantial disparity between adults living closer to screening institution and those who live farther regarding LCS use and among current smokers who were more likely to be screened. Our findings provide insights into targets for future interventions to promote lung cancer screening rates.

### Declarations

#### Ethical Approval

This study was approved by both University of Alabama at Birmingham and Morehouse School of Medicine

#### Transformative

I confirm that I understand journal Cancer Causes & Control is a transformative journal. When research is accepted for publication, there is a choice to publish using either immediate gold open access or the traditional publishing route.

#### Dual Publication

The results/data/figures in this manuscript have not been published elsewhere, nor are they under consideration by another publisher.

#### Authorship

I have read the Nature Portfolio journal policies on author responsibilities and submit this manuscript in accordance with those policies.

#### Third Party Material

All of the material is owned by the authors and/or no permissions are required.

#### Data Availability

Yes, I have research data to declare. Data will be made available upon request.

## Data Availability

All data produced in the present study are available upon reasonable request to the authors

